# Predictors of Premature Discontinuation of Opioid Use Disorder Treatment in the United States

**DOI:** 10.1101/2021.07.26.21261080

**Authors:** Celia Stafford, Wesley Marrero, Rebecca B. Naumann, Kristen Hassmiller Lich, Sarah Wakeman, Mohammad S. Jalali

## Abstract

Over the last few decades, opioid use disorder (OUD) and overdose have dramatically increased. Evidence shows that treatment for OUD, particularly medication for OUD, is highly effective; however, despite decreases in barriers to treatment, retention in OUD treatment remains a challenge. Therefore, understanding key risk factors for OUD treatment discontinuation remains a critical priority. We built a machine learning model using the Treatment Episode Data Set – Discharge (TEDS-D). Included were 2,446,710 treatment episodes for individuals in the U.S. discharged between January 1, 2015 and December 31, 2018 (the most recent available data). Exposures contain 32 potential risk factors, including treatment characteristics, substance use history, socioeconomic status, and demographic characteristics. Our findings show that the most influential risk factors include characteristics of treatment service setting, geographic region, primary source of payment, referral source, and health insurance status. Importantly, several factors previously reported as influential predictors, such as age, living situation, age of first substance use, race and ethnicity, and sex had far weaker predictive impacts. The influential factors identified in this study should be more closely explored to inform targeted interventions and improve future models of care.

## Introduction

Opioid-related deaths have risen dramatically since the 1990’s, with 49,860 deaths in the US in 2019 alone. With an estimated 1.6 million people suffering from an opioid use disorder (OUD),^1^ millions more lives are impacted beyond just victims of fatal overdose.^2,3^

Treatment for OUD, particularly medication for OUD (MOUD), is highly effective.^4^ Treatment success depends on adherence and retention in care.^5^ Research indicates that while there is variation by treatment type (e.g., MOUD or psychosocial treatment), OUD treatment of all kinds suffers from high rates of premature exit. In some cases, treatment discontinuation has been reported as high as 85%,^6^ although the average rate of exit hovers around 30%.^7^ Beyond the US; studies citing treatment discontinuity are numerous globally.^8,9^ Better retention in treatment requires an improved understanding of the characteristics and needs of target populations, and to date, the literature offers limited and conflicting evidence.

Two systematic reviews (published in 2013^10^ and 2020^7^) summarize several hundred studies on substance use disorder (SUD) treatment attrition. They show that literature on predictors of treatment retention, adherence, or discontinuation shares small sample sizes (e.g., median n=144 among studies included in the 2013 review), and limited scope and generalizability (e.g., faith-based treatment, or male populations only). Also, the reviewed studies considered few covariates, and primarily demographics (e.g.,^11–14^). Furthermore, where there is overlap on factors included across studies, the results are often conflicting. For example, 64 of the 122 studies included in the 2013 review examined the role of sex. Just 10 of those reported a statistically significant relationship, with five reporting male sex as a predictor of treatment discontinuation and five reporting female sex as a predictor of discontinuation. Less than 10% of studies investigated the relationship between retention and treatment specific factors such as the method, setting, and duration.^10^ In the 2020 review,^7^ a similar focus on age, sex, and education was evident. For example, 146 of the 151 studies examined the role of age, with studies generally concluding that age was not a significant predictor of premature treatment exit. The only participant characteristics associated with discontinuation were race, income, daily cigarettes smoked, and heroin and cocaine use. Since the publication of these systematic reviews, a 2021 study specific to retention in MOUD found that methamphetamine use, younger age, and homelessness were risk factors for treatment discontinuation.^15^

Despite the substantial body of literature on the opioid overdose crisis, the importance of treatment, and concerns about treatment retention,^16^ a large-scale analysis of demographic *and* contextual factors contributing to premature treatment exit has not been conducted. To address this gap, we developed a machine learning model based on millions of treatment episodes with a large holistic set of predictors. Our objective was to examine which factors best predict treatment attrition. Illuminating specific factors from this rich, national data source can help pinpoint factors that should be more closely explored to inform targeted interventions.

Additionally, though machine learning-based predictive models have been widely used in medicine,^17^ there is no big-data-based model utilized for understanding premature OUD treatment exit. We develop the first model of its kind and identify key factors and their relationship with premature OUD treatment exit.

## Methods

### Study Population

The cohort consisted of OUD treatment episodes included in the Treatment Episode Data Set-Discharge (TEDS-D)^18^—a national data system that contains records of individuals ages 12 and above derived from SUD treatment facilities. The national system includes facilities that receive any federal funding—varying by state, this can also include facilities such as private doctors’ offices.^19^ Annual data from 2015, until the most recent available year, 2018, were combined. Years prior to 2015 were excluded as they collected less data. Included were episodes where individuals reported use of heroin, or other opioids. No other inclusion or exclusion criteria were used.

We followed best practice observational study reporting guidelines.^20^ Complete datasets, code, and results are available on GitHub (https://github.com/castaff/TEDS_Treatment_Attrition) for review and reproducibility.^17^

### Factors and Factor Creation

To predict treatment discontinuation, we included 23 factors from TEDS-D that were collected at admission and excluded factors collected at treatment discharge. Included in this analysis are demographics, frequency of substance use, routes of administration, self-reported substances of use, treatment history, source of referral, and planned treatment type (i.e., whether MOUD is planned in the patient’s treatment).

By combining and recoding existing factors, we also created several additional variables. The first reflects minimum reported age at which the individual began using substances.^21,22^ The second presents the maximum frequency of nonmedical opioid use. A third variable was created indicating heroin use, and the fourth indicated any injection drug use.^23^ Finally, four binary factors were created indicating whether individuals used substances falling into the broad categories of stimulants, hallucinogens, sedatives, or tranquilizers (Table S1).

In total, 32 factors were included. A table of variables excluded and the reasons for exclusion can be found in Table S1, as well as a complete list of the variables included and their definitions. Missing data were imputed using a random forest-based multiple imputation approach.^24^

### Outcome

We dichotomized the reason for discharge to “dropped out of treatment” vs. all other reasons for treatment discharge. “Dropped out of treatment” includes clients who exited treatment for unknown reasons as well those who left against professional advice or were lost to follow up and discharged administratively. The outcome is non-missing for all treatment episodes.

### Machine Learning Analysis

We conducted a classification analysis, using a tree-based approach. With decision trees, what is made up for in interpretability is lost in accuracy as there can be large differences between trees based on changes in the data sampled for model training.^25^ This variance can be improved by building a “forest” of many trees. Differentiating it from bagged trees, a random forest selects parameters for each split from a random selection of *m* variables. This reduces correlation among individual trees. The majority vote is then taken of the predicted class from all trees.^26^

The data were divided into a training and a testing set, using a 75% and 25% split. We performed 10-fold cross validation on the 75% training set, and a final model was fit on the complete training set and performance assessed on the testing set. Missing data were imputed on the training set and the testing set independently to avoid data leakage.^24^ Additionally, models were fit based upon data stratified by year, as well as only on complete records (without imputation). Model performance was assessed with area under the receiver operating curve (AUC) and accuracy. In addition, permutation variable importance, which computes the decrease in model performance when a given predictor is shuffled, was reported. Finally, partial dependence plots were created for the top five influential variables. These plots depict the marginal effect a variable has on the predicted outcome of a given machine learning model.^27^ All analyses were performed using R statistical programming language version 3.3, using the tidymodels, ranger, vip, broom, pdp and missForest packages. Data preprocessing was conducted with dplyr.

## Results

Figure 1 presents an overview of study data. After excluding variables not collected at treatment admission and episodes without reported opioid use, 2,446,710 episodes were included in the analysis. Of these treatment episodes, 321,735 contained complete data. Most variables had missing values on less than 10% of the records, with just health insurance status, primary source of payment for treatment, days waited before entering treatment, primary source of income, and marriage status missing more than 20%. Overall, about 90% of observations were missing five or fewer variables. A complete summary of missingness can be found in Table S2.

**Figure 1:**
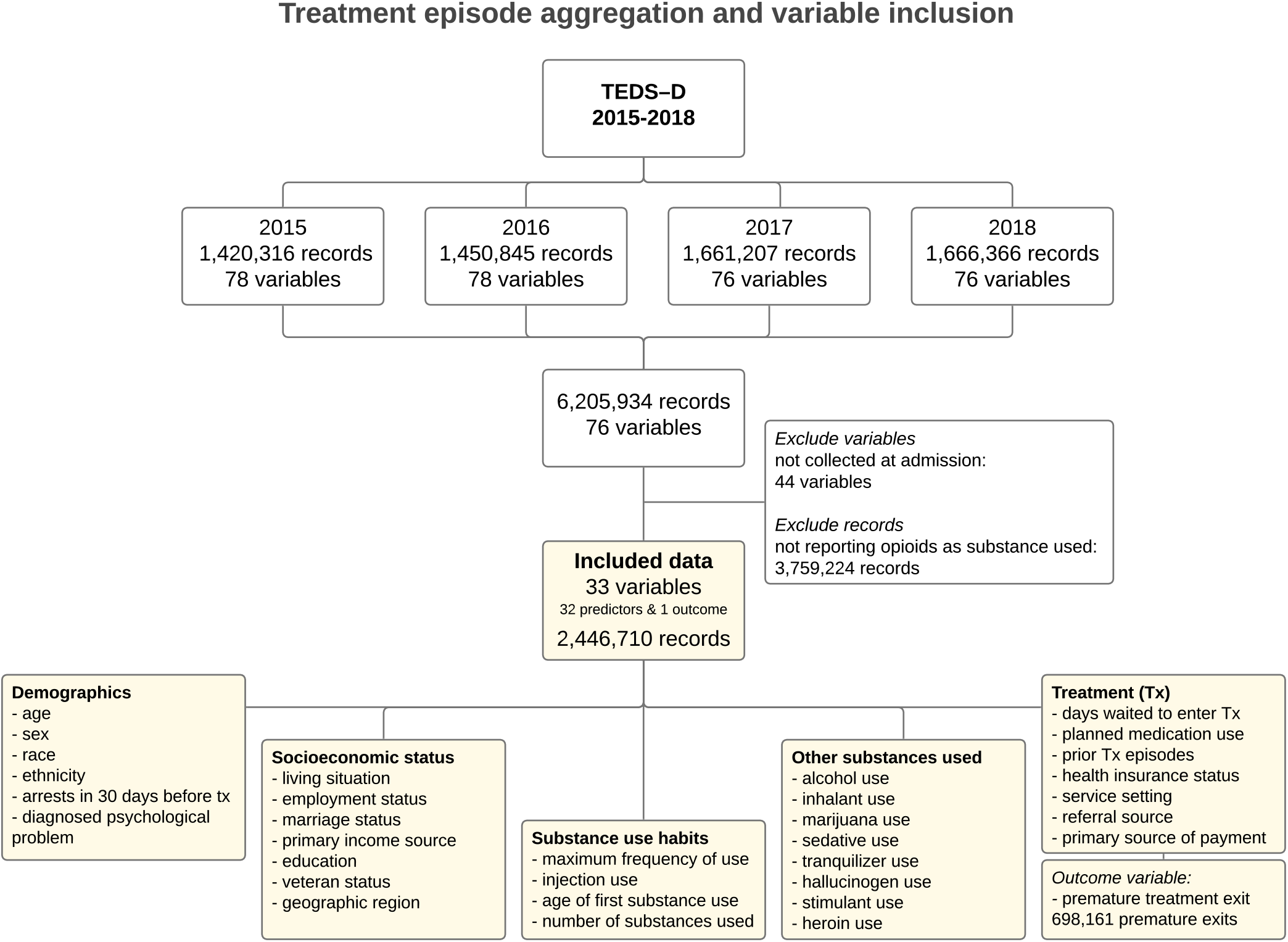
Episode and variable inclusion and exclusion

### Descriptive Results

Table S3 presents a full listing of the characteristics of individual treatment episodes, stratified by premature treatment exit status. The vast majority (76%) of opioid-related treatment episodes were among white individuals. Of all opioid-related episodes, stimulants were the most often co-occurring substance reported (37%), followed by cannabis (21%) and alcohol (21%). A total of 698,161 (28.5%) opioid-related treatment episodes resulted in treatment discontinuation. The most represented age group among all episodes was 25-34, which accounted for 43% of all episodes and 43% of the episodes resulting in premature exit. Of episodes resulting in premature exit, 27% had no reported prior treatment episodes. Among episodes in which individuals discontinued treatment, 34% had planned to use MOUD at baseline, while 24% of those who did not exit treatment prematurely planned to use MOUD at baseline.

### Random Forest

We used 1,835,033 treatment episodes for training and 611,677 for testing (see Figure S1). The random forest classifier achieved a mean AUC of 75% with a standard error of 0.002 across the 10 folds cross-validation. On the unseen-by-the-model testing set, the random forest achieved an accuracy of 72% with an AUC of 70%.

Of greater relevance to treatment decisions is the relative importance of included factors in predicting premature treatment exit. A full ranking of important factors can be seen in Figure 2. The most influential predictor was service setting (e.g., inpatient, ambulatory, detox). Its exclusion from the model decreased accuracy by almost 4%. Besides service setting, geographic region, referral source, primary source of payment for treatment, and health insurance status each produced an accuracy decrease greater than 1.5%. Interestingly, low on the list of importance were reported heroin use, injection use, age, race, and ethnicity. Variable importance remained largely unchanged when the model was fit on individual years (see Figure S2).

**Figure 2:**
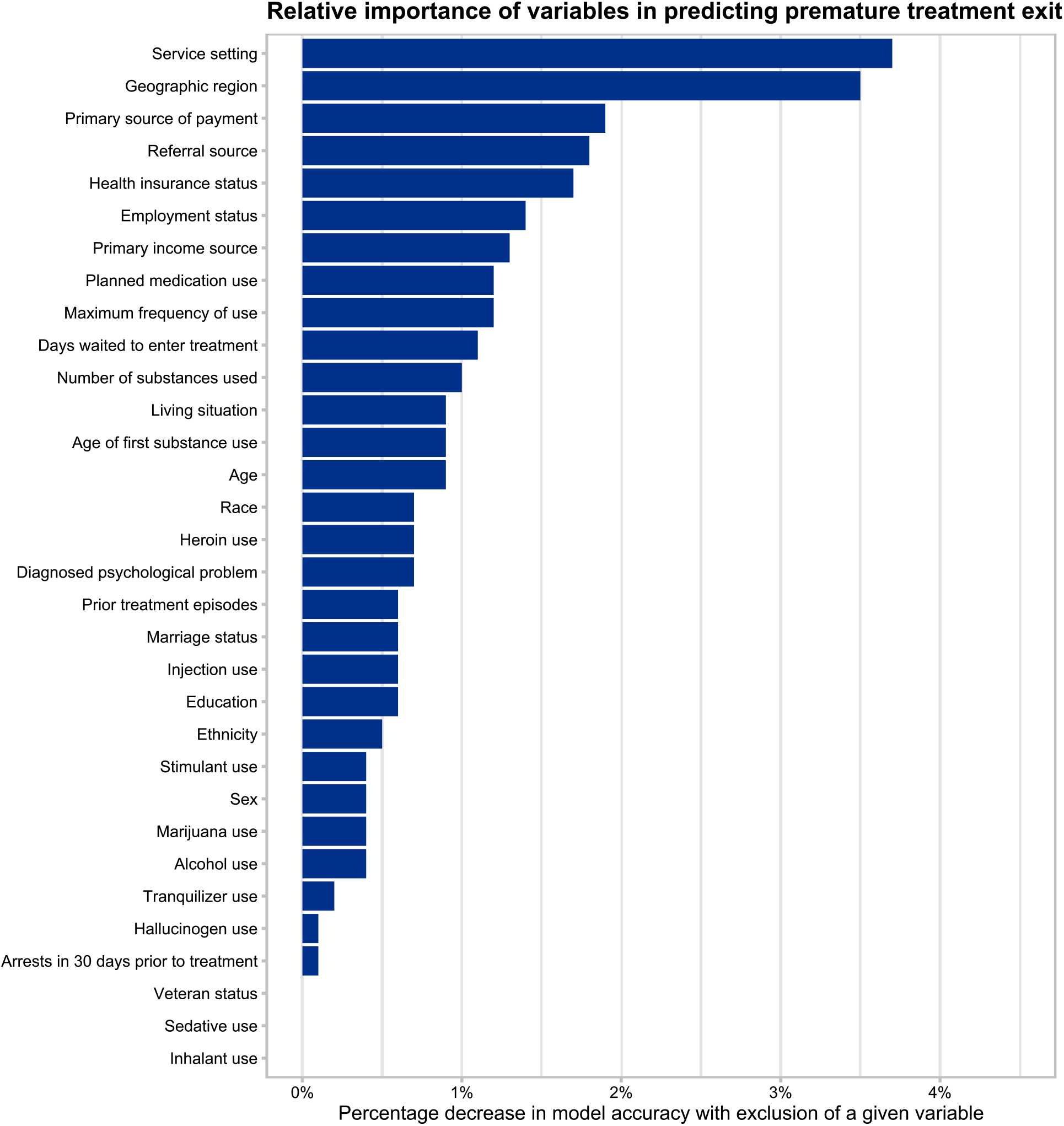
Variable importance of the predictors of premature treatment exit

Figure 3 shows the marginal impact of different levels of each of the five most important variables on treatment dropout with partial dependence plots—service setting of 24hr freestanding residential detox, southern geographic region, Medicaid source of payment and insurance type, and criminal justice treatment referral were associated with the largest decreases in treatment retention.

**Figure 3:**
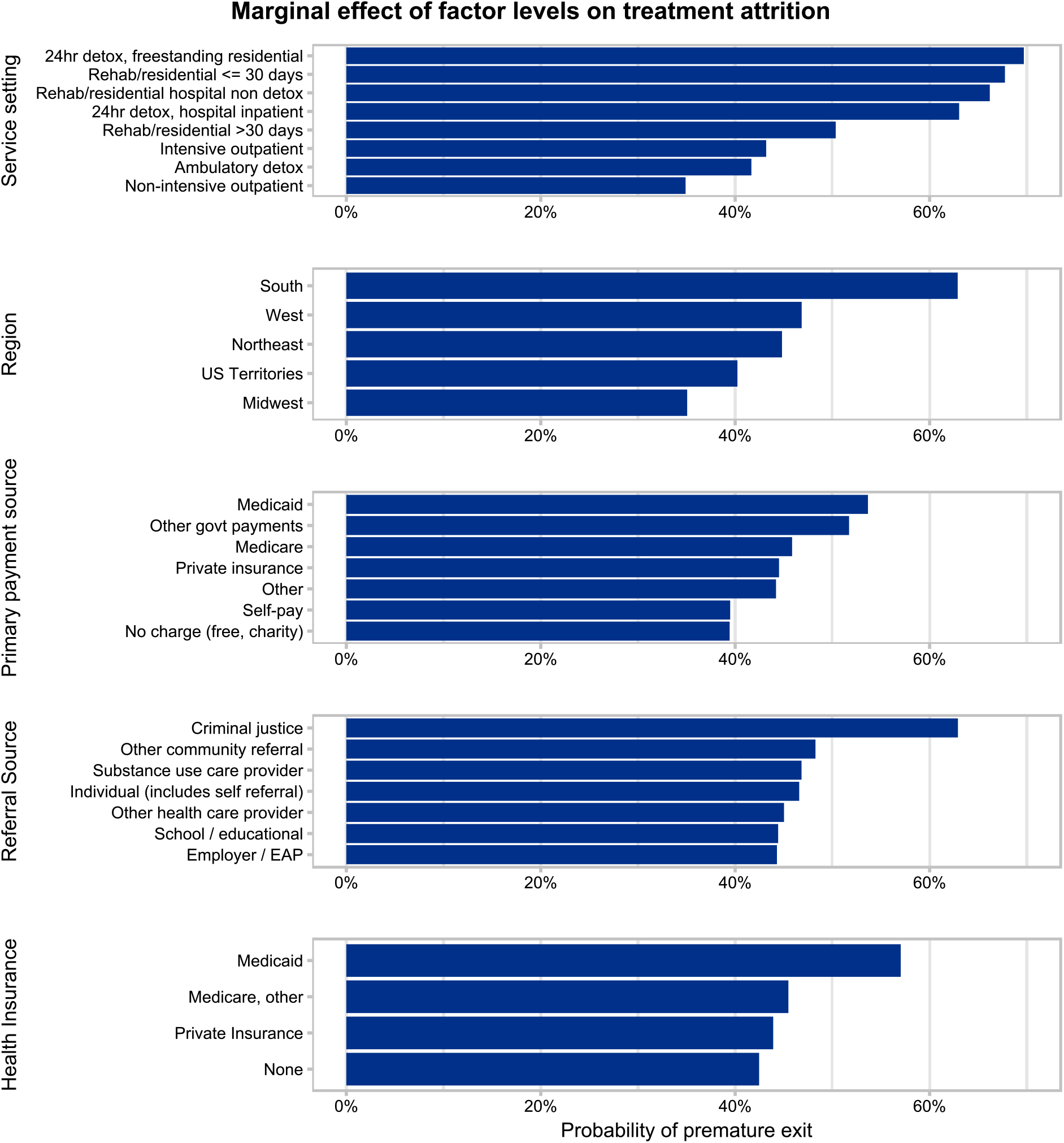
Partial dependence plots of most influential variables

## Discussion

We used a machine learning model to predict premature OUD treatment exit using treatment episode data from TEDS-D. With 32 predictors spanning demographics, substance use habits, and treatment information, we built a model identifying factors most influential in determining whether an individual will exit treatment prior to completion. We found that treatment setting, geographical region, health insurance status, treatment referral source, and primary source of payment were the strongest predictors.

While we found that service setting was the most important predictor of attrition, only 3% of papers reviewed by Brorson et al did.^10^ Service setting options captured in TEDS-D include detox, short- and long-term inpatient care, and different forms of ambulatory care, each with differing implications for transportation, finances, and other factors. Though longer term and/or ambulatory care-based treatment modalities offer more chances to lose patients in the process of care, we found that patients are less likely to prematurely exit these. Certain treatments, such as MOUD, lead to better outcomes,^4, 555^ however there is little research showing that detox or residential care improves outcomes for individuals with OUD. Coupled with high attrition rates, it is clear that this is a costly and ineffective “revolving door” treatment model that bears re-evaluation. Thought should be dedicated to expanding long-term outpatient care potentially coupled with affordable housing rather than further investment in costly inpatient models.

Another notable feature is geographic region. Figure 3 highlights a meaningful difference in attrition across regions, and it is noticeable that exclusion of this variable from the model precipitates a drop in accuracy of over 3.5%. As shown in the second panel of Figure 3, likelihood of premature exit was highest in the South. Research indicates that this heterogeneity may exist due to infrastructural factors related to treatment access, social factors like stigma, and socioeconomic factors such as likelihood of working a manual labor job; a lot of this heterogeneity remains causally unexplained and is subject to more investigation.^28^

While socio-economic status and income are occasionally tested,^10^ our model is the first to include the primary source of payment for treatment. We find it to be the third most influential factor with highest rates of premature treatment exit in individuals paying with Medicare, Medicaid, and with other government payments. This connection bears further research – it is possible that programs accepting public payers differ from those accepting commercial insurance, or there could be ties to socioeconomic status and potential multicollinearity with social determinants of health or employment status. Simply knowing risk of treatment attrition is high among Medicaid enrollees provides an intervention opportunity.

Related to the primary source of payment, we also surfaced health insurance status as an important predictor. Medicaid stands out as a predictor of treatment attrition. Research indicates that Medicaid beneficiaries have a high prevalence of comorbidities and barriers to healthcare, including affordable and accessible transportation, that may contribute to this attrition.^29^ More recently, state Medicaid programs have attempted to remove insurance-based barriers (e.g., prior authorizations) to MOUD initiation; however, additional work to support treatment retention remains a clear need.

Referral source was also an important predictive factor, with referral from criminal legal settings associated with increased probability of premature treatment exit. Research indicates that 15% of deaths following release from prison are related to opioids and decreased tolerance thereof, and up to 65% of the US prison population may have an active substance use disorder.^30^ The Substance Abuse and Mental Health Services Administration and others have increased resources to support access to MOUD in prisons, and post-release; however, additional supports are needed. It is also apparent that coerced treatment, i.e., treatment mandated by the courts, may not be an effective strategy.^31^

With interestingly low impact on model accuracy were age and age of first use. Although age of first substance use has not been investigated in this context, there is evidence of its tie to development of OUD,^21^ and eventual admission to treatment.^22^ Most studies assess the impact of age on treatment exit, but just 36% of the studies investigating age found a significant relationship. Of these, 88% linked younger age to increased risk of premature treatment exit.^7,10^ Additional research on how substance use trajectories influence treatment initiation and retention trends is needed, as well as how treatment retention supports may differ across the life span.

In contrast with the many small cohort and retrospective studies on premature treatment discontinuation, this analysis utilized records from millions of substance use treatment episodes. While these data have been widely studied, research has focused on trends of substance use over time.^32,33^ We leveraged these data to study predictors of treatment discontinuation, taking advantage of a rich set of covariates and the 2,446,710 treatment episodes. This provided a larger sample size and the ability to explore a wider range of risk factors than any other study on the subject. Also differentiating this study, we utilized a supervised machine learning approach. This method offers a distinct benefit over previous analyses by considering possible interactions and multicollinearity among factors. Finally, random forest models are helpful in assessing nonlinearities which is challenging to parse out with classic regression-based models.

This study is subject to several limitations. First, TEDS-D is an observational dataset that relies on submission by individual states. Depending on year, several states do not submit data and are not included in totals. Nonresponse may be associated with higher rates of treatment discontinuation due to less treatment funding or substance use support which may lead to underestimates of rates of treatment exit. However, we would not expect this to alter the predictive relationships explored in our models. Second, TEDS-D has a minimum set of data elements that states are required to report on, including demographic and substance use factors. Factors outside of this minimum requirement can be missing. These missing data were imputed using a random forest and though the efficacy of this method has been described in detail (e.g., ^34,35^), imputation is not a perfect solution. We report our results on complete records (without imputation) in Figure S3 and the findings are overall similar to those with imputation.

TEDS-D also includes only facilities that are state licensed or certified. Because of differences in individual state policies, some states include private doctors’ offices and other private clinics, and some do not. While it is the most complete survey of treatment facilities, TEDS-D is an undercount both because of non-reporting and excluded private facilities. Additionally, there could be bias around the kinds of people that exit state-sponsored facilities vs those that might exit treatment prematurely at a private doctor’s office.

Reasons for treatment discharge not considered to be premature treatment discontinuation initiated by the patient included completion, termination by the facility and transfer to another facility or treatment program; a future analysis could consider any premature exit, whether initiated by patient or facility, as a failure. Finally, there is considerable variation in response to opioid use treatment. Because of this, there is great need to identify patients who will not respond well or are more likely to exit treatment prematurely. Although we identified key predictors of premature treatment exit, we cannot conclude that these are causal relationships. Still, these serve as important areas for future research to further explore specific causal mechanisms. Understanding the dynamics surrounding these key factors holds important clinical relevance for future treatment decisions and models of care.

## Conclusion

We demonstrated that a predictive model for premature OUD treatment exit can be constructed using a dataset of US adults receiving treatment for OUD at state-affiliated treatment facilities. Our results may help address varying likelihoods of treatment attrition across patients and modalities of treatment. The combination of effective treatment interventions with data on an individual’s risk level can help channel resources toward targeted mechanisms of attrition for specific patients.

## Data Availability

All data and analysis details are reported in the article.

https://github.com/castaff/TEDS_Treatment_Attrition

## Ethical approval

Not needed.

## Supplementary Document for

### Supplementary Figures

**Figure S1.**
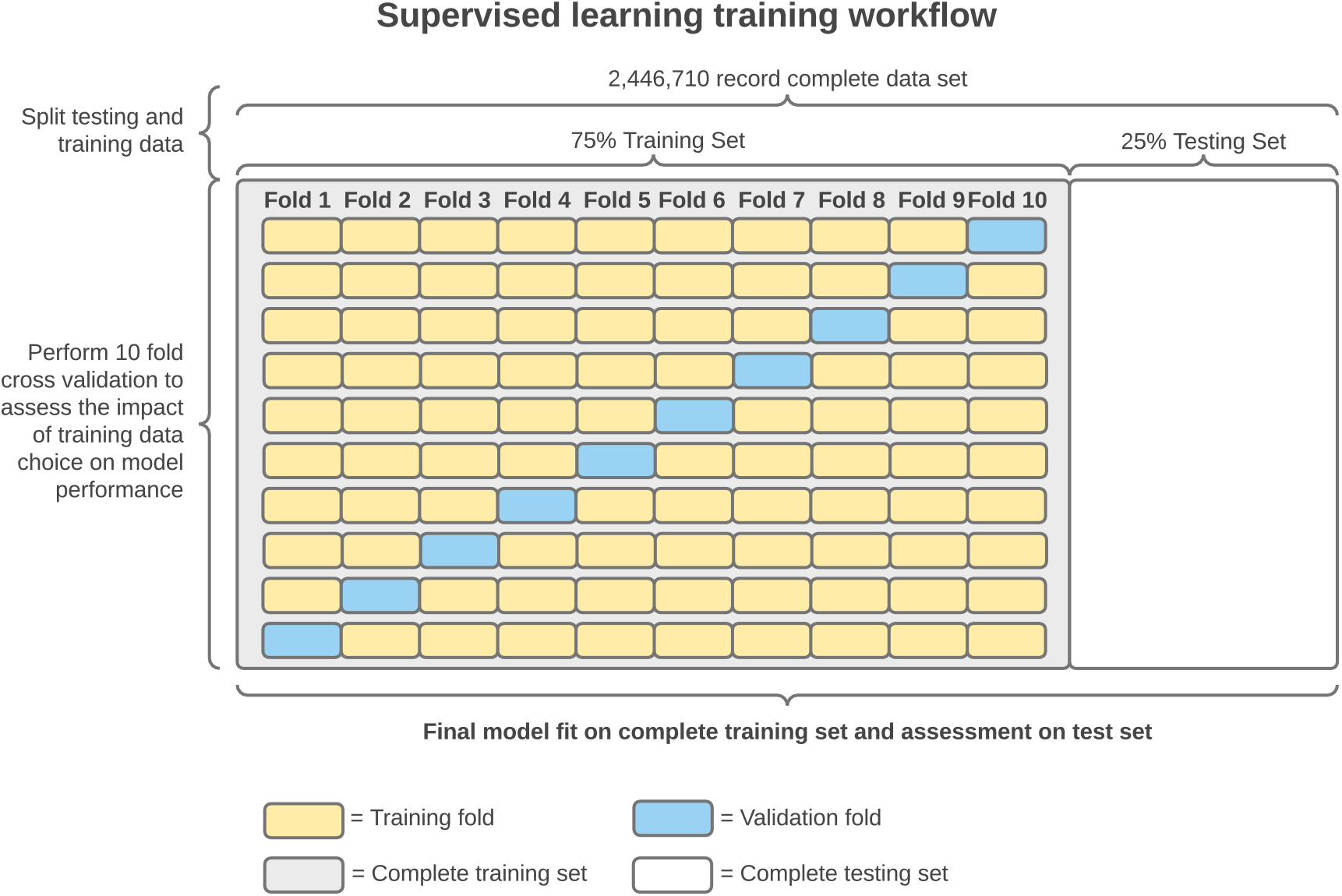
Data partitioning for cross validation

**Figure S2.**
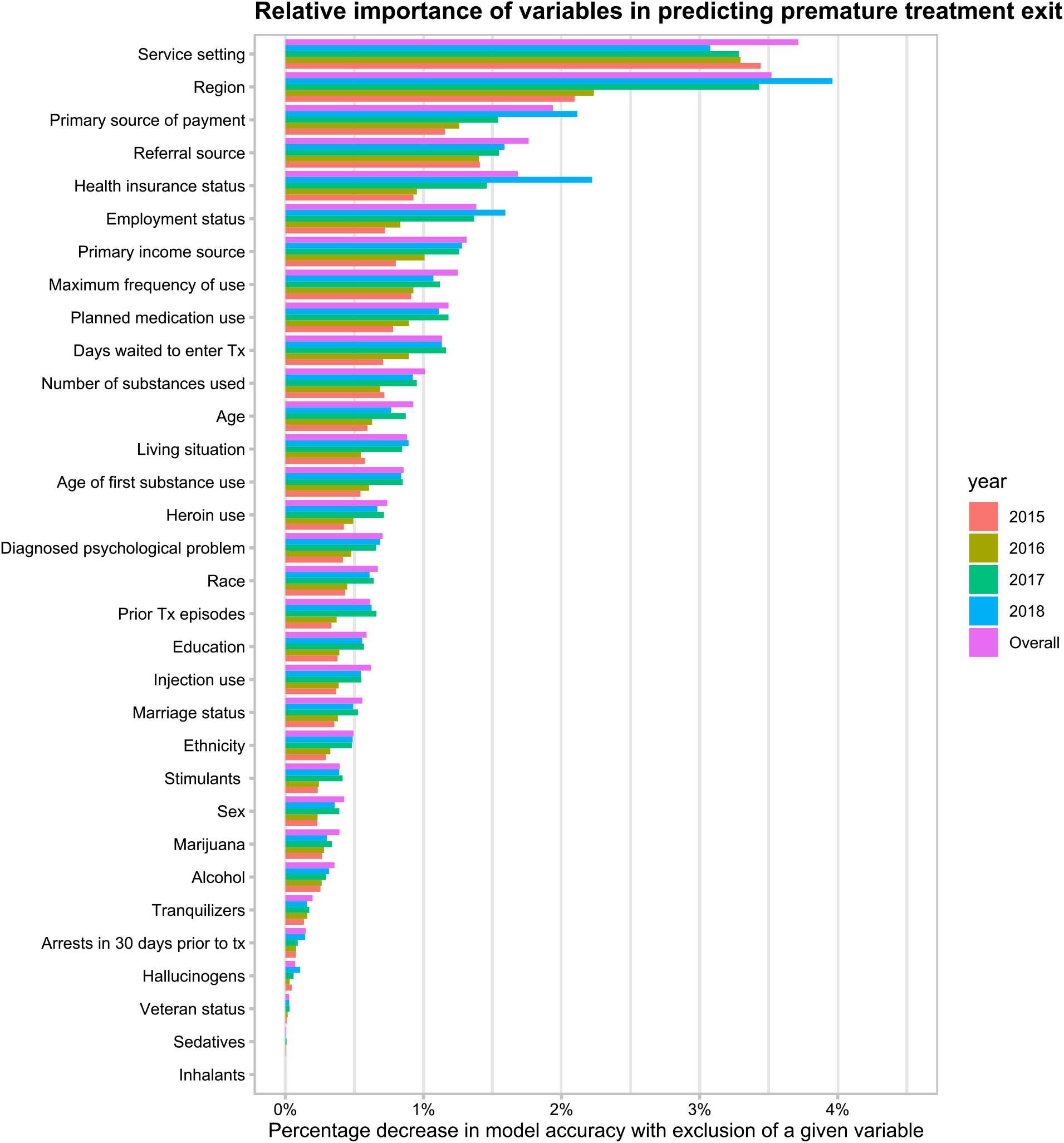
Variable importance stratified by year

**Figure S3.**
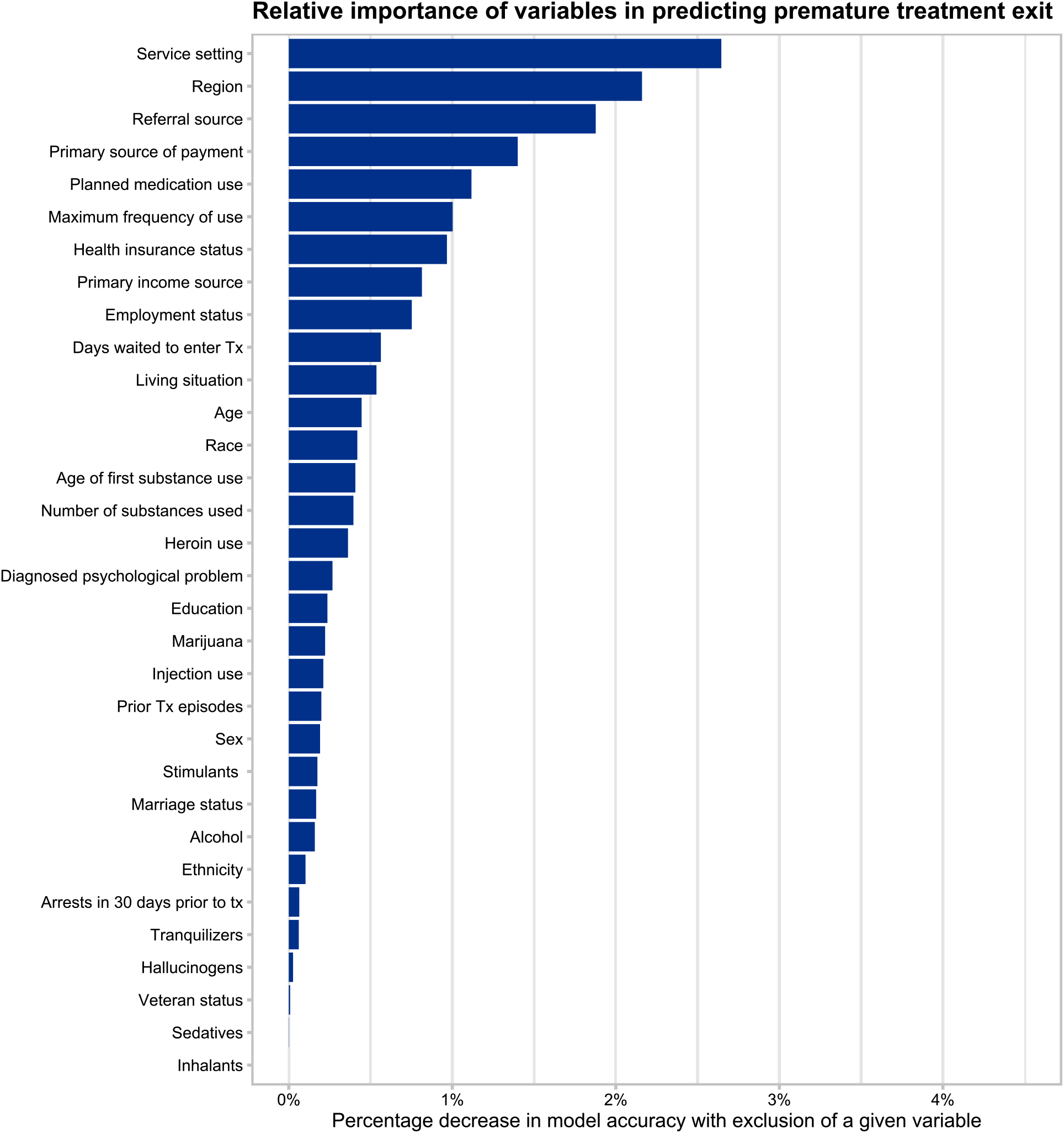
Variable importance of complete data records without data imputation

### Supplementary Tables

**Table S1.** TEDS-D variable exclusion

| Variable Label in TEDS | Description | Reason for Exclusion | Feature Adjustment |
| --- | --- | --- | --- |
| CASEID | ID Number | Not relevant for analysis |  |
| DETCRIM | Detailed criminal justice referral | Relevant only for a small percentage of individuals |  |
| DETNLF | Detailed not in labor force reason | Relevant only for a small percentage of individuals |  |
| FREQ1 | Frequency of use of first reported substance |  | Recoded into max frequency of opioid use |
| FRSTUSE2 | Age of first use of second reported substance |  | Recoded into age of first substance use |
| FREQ_ATND_SELF_HELP | Frequency of attendance of self help | Relevant only for certain episodes |  |
| SUB1_D | Primary substance used at discharge | Only variables collected at admission were considered |  |
| LIVARAG_D | Living arrangements at discharge | Only variables collected at admission were considered |  |
| FREQ3_D | Frequency of use of third reported substance at discharge | Only variables collected at admission were considered |  |
| METHFLG | Methadone flag |  | Recoded into opioid flag variable |
| MTHAMFLG | Methamphetamine flag |  | Recoded into stimulant flag variable |
| TRNQFLG | Tranquilizer flag |  | Recoded into tranquilizer flag variable |
| OTCFLG | Over the counter substance flag | Severe class imbalance - only applies to less than a half of a percent of treatment episodes |  |
| DIVISION | Census division | Region was kept as the geographical factor |  |
| CBSA | CBSA Code | Large number of levels - creates bias in random forests |  |
| STFIPS | Census state FIPS code | Large number of levels - creates bias in random forests |  |
| SUB1 | Primary substance used |  | Accounted for with the various flag variables |
| FRSTUSE1 | Age of first use of first reported substance |  | Recoded into age of first substance use |
| ROUTE3 | Route of administration of third reported substance |  | Recoded into injection use flag |
| DSMCRIT | DSM Diagnoses | Over 90% missing |  |
| SUB2_D | Secondary substance used at discharge | Only variables collected at admission were considered |  |
| DETNLF_D | Detailed not in labor force reason at discharge | Only variables collected at admission were considered |  |
| FREQ_ATND_SELF_HELP_D | Frequency of attendance of self help at discharge | Only variables collected at admission were considered |  |
| COKEFLG | Cocaine flag |  | Recoded into stimulant flag variable |
| OPSYNFLG | Opioid and other synthetics flag |  | Recoded into opioid flag variable |
| AMPHFLG | Amphetamine flag |  | Recoded into stimulant flag variable |
| BARBFLG | Barbiturate flag |  | Recoded into sedative flag variable |
| OTHERFLG | Other drug flag |  | Recoded into hallucinogen flag variables |
| SUB2 | Secondary substance used |  | Accounted for with the various flag variables |
| PREG | Pregnancy status | Relevant only for a small percentage of individuals |  |
| ROUTE2 | Route of administration of second reported substance |  | Recoded into injection use flag |
| FREQ3 | Freq of administration of third reported substance |  | Recoded into max frequency of opioid use |
| SERVICES_D | Service setting at discharge | Only variables collected at admission were considered |  |
| SUB3_D | Tertiary substance of use at discharge | Only variables collected at admission were considered |  |
| FREQ1_D | Frequency of use of primary substance at discharge | Only variables collected at admission were considered |  |
| LOS | Length of stay in treatment | Only variables collected at admission were considered |  |
| PCPFLG | PCP flag |  | Recoded into hallucinogen flag variable |
| STIMFLG | Stimulant flag |  | Recoded into stimulant flag variable |
| SEDHPFLG | Sedative flag |  | Recoded into sedative flag variable |
| NUMSUBS | Number of substances used |  | Included |
| ALCDRUG | Alcohol and drug or just alcohol or drug use |  | Accounted for with the various flag variables |
| SUB3 | Third substance used |  | Accounted for with the various flag variables |
| ROUTE1 | Route of administration for primary substance used |  | Recoded into injection use flag |
| FREQ2 | Frequency of use for secondary substance used |  | Recoded into max frequency of opioid use |
| FRSTUSE3 | Age of first use of tertiary substance used |  | Recoded into age of first substance use |
| REASON | Reason for discharge |  | Recoded into binary outcome variable |
| EMPLOY_D | Employment status at discharge | Only variables collected at admission were considered |  |
| FREQ2_D | Frequency of use of secondary substance at discharge | Only variables collected at admission were considered |  |
| ARRESTS_D | Arrests at discharge | Only variables collected at admission were considered |  |
| HERFLG | Heroin flag |  | Recoded into opioid flag variable |
| HALLFLG | Hallucinogen flag |  | Recoded into hallucinogen flag variable |
| BENZFLG | Benzodiazepine flag |  | Recoded into tranquilizer flag variable |
| IDU | Injection drug use | Indicates primary injection use, a variable indicating any injection was included in this analysis instead |  |
| YEAR | Year |  | Included only for individual year analysis, not used as a predictor |

**Table S2.**
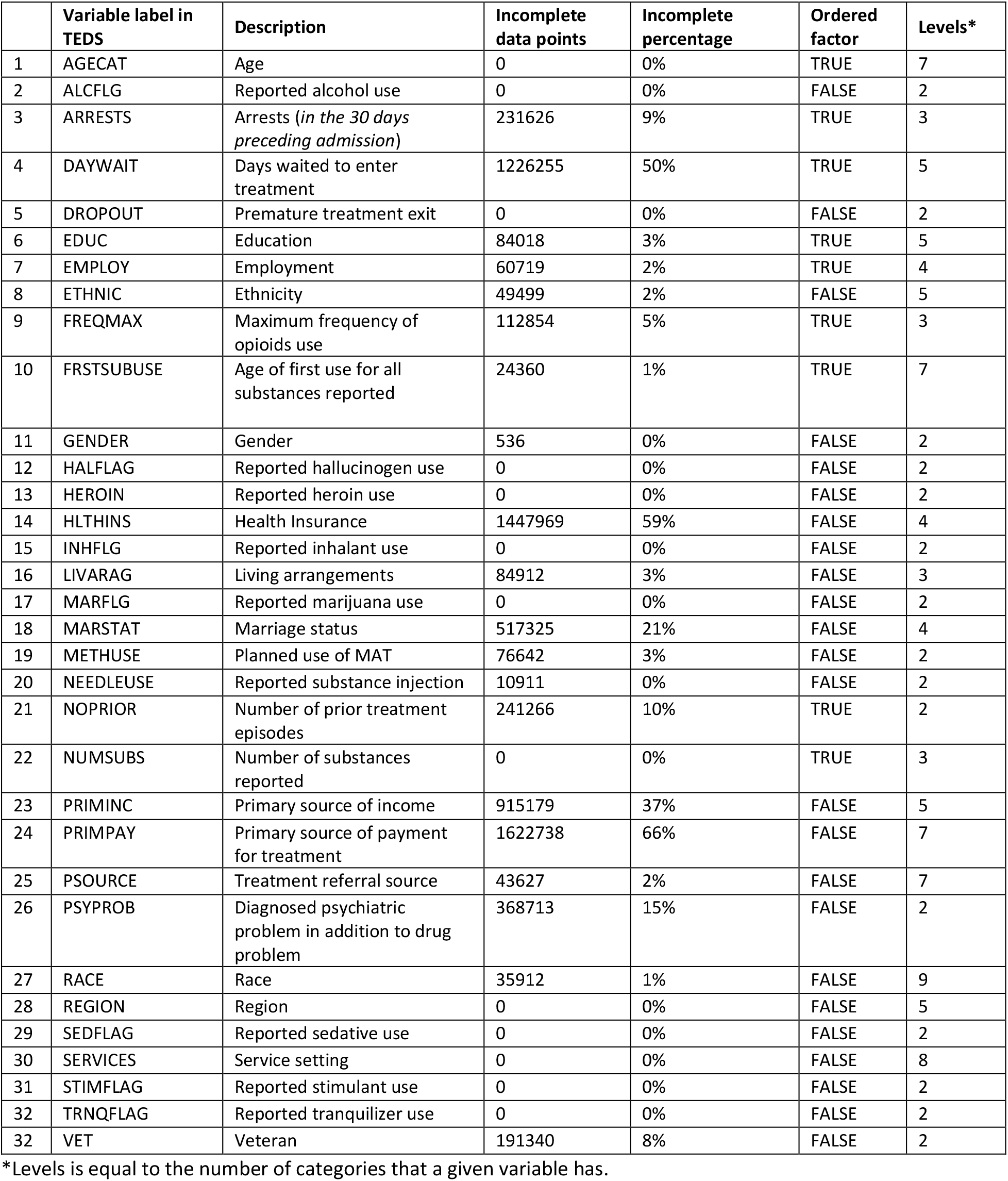
Missing data summary

|  | Variable label in TEDS | Description | Incomplete data points | Incomplete percentage | Ordered factor | Levels* |
| --- | --- | --- | --- | --- | --- | --- |
| 1 | AGECAT | Age | 0 | 0% | TRUE | 7 |
| 2 | ALCFLG | Reported alcohol use | 0 | 0% | FALSE | 2 |
| 3 | ARRESTS | Arrests (*in the 30 days preceding admission*) | 231626 | 9% | TRUE | 3 |
| 4 | DAYWAIT | Days waited to enter treatment | 1226255 | 50% | TRUE | 5 |
| 5 | DROPOUT | Premature treatment exit | 0 | 0% | FALSE | 2 |
| 6 | EDUC | Education | 84018 | 3% | TRUE | 5 |
| 7 | EMPLOY | Employment | 60719 | 2% | TRUE | 4 |
| 8 | ETHNIC | Ethnicity | 49499 | 2% | FALSE | 5 |
| 9 | FREQMAX | Maximum frequency of opioids use | 112854 | 5% | TRUE | 3 |
| 10 | FRSTSUBUSE | Age of first use for all substances reported | 24360 | 1% | TRUE | 7 |
| 11 | GENDER | Gender | 536 | 0% | FALSE | 2 |
| 12 | HALFLAG | Reported hallucinogen use | 0 | 0% | FALSE | 2 |
| 13 | HEROIN | Reported heroin use | 0 | 0% | FALSE | 2 |
| 14 | HLTHINS | Health Insurance | 1447969 | 59% | FALSE | 4 |
| 15 | INHFLG | Reported inhalant use | 0 | 0% | FALSE | 2 |
| 16 | LIVARAG | Living arrangements | 84912 | 3% | FALSE | 3 |
| 17 | MARFLG | Reported marijuana use | 0 | 0% | FALSE | 2 |
| 18 | MARSTAT | Marriage status | 517325 | 21% | FALSE | 4 |
| 19 | METHUSE | Planned use of MAT | 76642 | 3% | FALSE | 2 |
| 20 | NEEDLEUSE | Reported substance injection | 10911 | 0% | FALSE | 2 |
| 21 | NOPRIOR | Number of prior treatment episodes | 241266 | 10% | TRUE | 2 |
| 22 | NUMSUBS | Number of substances reported | 0 | 0% | TRUE | 3 |
| 23 | PRIMINC | Primary source of income | 915179 | 37% | FALSE | 5 |
| 24 | PRIMPAY | Primary source of payment for treatment | 1622738 | 66% | FALSE | 7 |
| 25 | PSOURCE | Treatment referral source | 43627 | 2% | FALSE | 7 |
| 26 | PSYPROB | Diagnosed psychiatric problem in addition to drug problem | 368713 | 15% | FALSE | 2 |
| 27 | RACE | Race | 35912 | 1% | FALSE | 9 |
| 28 | REGION | Region | 0 | 0% | FALSE | 5 |
| 29 | SEDFLAG | Reported sedative use | 0 | 0% | FALSE | 2 |
| 30 | SERVICES | Service setting | 0 | 0% | FALSE | 8 |
| 31 | STIMFLAG | Reported stimulant use | 0 | 0% | FALSE | 2 |
| 32 | TRNQFLAG | Reported tranquilizer use | 0 | 0% | FALSE | 2 |
| 32 | VET | Veteran | 191340 | 8% | FALSE | 2 |

\*Levels is equal to the number of categories that a given variable has.

**Table S3.** Individual characteristics stratified by premature treatment exit status

| Variable | Levels | Premature Treatment Exit (%) |  |
| --- | --- | --- | --- |
|  |  | No | Yes |
|  |  | N=1,748,549 | N=698,161 |
| Demographics |  |  |  |
| Age | 12-17 | 0.1 | 0 |
|  | 18-24 | 14.7 | 15.2 |
|  | 25-34 | 43.6 | 42.9 |
|  | 35-44 | 22.1 | 21.8 |
|  | 45-54 | 13.2 | 13.5 |
|  | 55-64 | 5.7 | 5.8 |
|  | 65+ | 0.6 | 0.7 |
| Sex | Male | 62 | 62.1 |
|  | Female | 38 | 37.9 |
| Race | Alaska Native (Aleut, Eskimo, Indian) | 0.1 | 0.1 |
|  | American Indian (Other than Alaska Native) | 1.3 | 1.6 |
|  | Asian or Pacific Islander | 0.1 | 0 |
|  | Black or African American | 10.6 | 11.2 |
|  | White | 77 | 73.7 |
|  | Asian | 0.5 | 0.5 |
|  | Other single race | 7.8 | 9.9 |
|  | Two or more races | 2.3 | 2.6 |
|  | Native Hawaiian or other pacific islander | 0.3 | 0.3 |
| Ethnicity | Puerto Rican | 5.4 | 6.6 |
|  | Mexican | 2.5 | 3.7 |
|  | Cuban or other specific Hispanic | 2.3 | 2.7 |
|  | Not of Hispanic or Latino origin | 87.8 | 84.6 |
|  | Hispanic or Latino, origin not specified | 2 | 2.4 |
| Arrests in 30 days preceding admission | None | 90.2 | 92.4 |
|  | Once | 7.9 | 6.3 |
|  | Two or more times | 1.8 | 1.2 |
| Diagnosed psychiatric problem | No | 54.8 | 56.5 |
|  | Yes | 45.2 | 43.5 |
| Socioeconomic status |  |  |  |
| Living Arrangements | Homeless | 16.3 | 15.3 |
|  | Dependent Living | 17.9 | 15.5 |
|  | Independent Living | 65.8 | 69.2 |
| Employment Status | Full-Time | 12.4 | 12.6 |
|  | Part-Time | 6.3 | 6.6 |
|  | Unemployed | 42.2 | 40.4 |
|  | Not in labor force | 39.1 | 40.4 |
| Marriage Status | Never Married | 64.4 | 64.6 |
|  | Now Married | 13.9 | 14.7 |
|  | Separated | 7.1 | 6.9 |
|  | Divorced, Widowed | 14.5 | 13.8 |
| Primary source of income | Wages/Salary | 21.3 | 21.6 |
|  | Public Assistance | 12.3 | 14.2 |
|  | Retirement/Pension, Disability | 8.7 | 9.5 |
|  | Other | 19 | 19.6 |
|  | None | 38.6 | 35.1 |
| Education | 8 years or less | 6.1 | 7 |
|  | 9-11 years | 19.3 | 20.7 |
|  | 12 (GED) | 48.3 | 47.7 |
|  |  | No | Yes |
|  |  | N=1,748,549 | N=698,161 |
|  | 13-15 | 21.1 | 20.1 |
|  | 16 or more | 5.2 | 4.4 |
| Veteran | No | 97.4 | 97.6 |
|  | Yes | 2.6 | 2.4 |
| Region | US Jurisdiction/Territory | 0.1 | 0.3 |
|  | Northeast | 43.2 | 45.5 |
|  | Midwest | 15.1 | 19.3 |
|  | South | 26.5 | 15.8 |
|  | West | 15.2 | 19.1 |
| **Substance use habits** |  |  |  |
| Maximum frequency of opioid use | No use in past month | 26.2 | 22.3 |
|  | Some use | 19.1 | 19.2 |
|  | Daily Use | 54.6 | 58.5 |
| Injection use | No | 45.9 | 44.8 |
|  | Yes | 54.1 | 55.2 |
| Age of first use for all substances reported | 11 years and younger | 6.1 | 6.2 |
|  | 12-14 | 21.7 | 21.5 |
|  | 15-17 | 26.1 | 25.7 |
|  | 18-20 | 17.8 | 17.9 |
|  | 21-24 | 11 | 11.1 |
|  | 25-29 | 8.3 | 8.5 |
|  | 30+ | 9 | 9.2 |
| Number of substances reported | 1 | 28.3 | 30.6 |
|  | 2 | 36.8 | 37 |
|  | 3 | 34.9 | 32.4 |
| **Other substances used** |  |  |  |
| Alcohol Use | No | 78.2 | 80.3 |
|  | Yes | 21.8 | 19.7 |
| Inhalant Use | No | 99.9 | 100 |
|  | Yes | 0.1 | 0 |
| Marijuana Use | No | 78.8 | 78.1 |
|  | Yes | 21.2 | 21.9 |
| Reported Sedative Use | No | 99.2 | 99.3 |
|  | Yes | 0.8 | 0.7 |
| Reported Tranquilizer Use | No | 88.4 | 89.9 |
|  | Yes | 11.6 | 10.1 |
| Reported Hallucinogen Use | No | 96.7 | 97.4 |
|  | Yes | 3.3 | 2.6 |
| Reported Stimulant Use | No | 62.7 | 63.5 |
|  | Yes | 37.3 | 36.5 |
| Heroin use | No | 74.2 | 25.8 |
|  | Yes | 70.5 | 29.5 |
| **Treatment characteristics** |  |  |  |
| Days waited to enter treatment | 0 | 50.4 | 48 |
|  | 1-7 | 30 | 31.9 |
|  | 8-14 | 8 | 8.6 |
|  | 15-30 | 6.7 | 6.7 |
|  | 31+ | 5 | 4.8 |
| Planned Use of MAT | No | 75.8 | 65.7 |
|  |  | No | Yes |
|  |  | N=1,748,549 | N=698,161 |
|  | Yes | 24.2 | 34.3 |
| Prior Treatment Experience | No prior treatment episodes | 29.3 | 27.3 |
|  | One or more prior treatment episodes | 70.7 | 72.7 |
| Health insurance status | Private Insurance, Blue Cross Blue Shield, HMO | 6.9 | 7.1 |
|  | Medicaid | 56.7 | 49.9 |
|  | Medicare, Other (Tricare, CHAMPUS) | 11.3 | 12.3 |
|  | None | 25.1 | 30.7 |
| Service Setting | Detox, 24 hr, hospital inpatient | 3.9 | 2.8 |
|  | Detox, 24 hr, freestanding residential | 22.8 | 18 |
|  | Rehab/residential, hospital (non-detox) | 0.2 | 0.2 |
|  | Rehab/residential, short term (30 days or fewer) | 13.2 | 8.7 |
|  | Rehab/residential, long term (more than 30 days) | 8.1 | 7.1 |
|  | Ambulatory, intensive outpatient | 12.8 | 11.8 |
|  | Ambulatory, non-intensive outpatient | 37.6 | 49.7 |
|  | Ambulatory, detoxification | 1.3 | 1.7 |
| Treatment Referral Source | Individuals (Includes Self-Referral) | 50.5 | 57.5 |
|  | Alcohol/Drug Use Care Provider | 14 | 13.1 |
|  | Other Health Care Provider | 6.3 | 6.5 |
|  | School (Educational) | 0.1 | 0.1 |
|  | Employer (EAP) | 0.2 | 0.2 |
|  | Other Community Referral | 8.7 | 8.2 |
|  | Court/Criminal Justice Referral/ DUI/ DWI | 20.2 | 14.4 |
| Primary source of payment for treatment | Self-Pay | 7.1 | 8.2 |
|  | Private Insurance, Blue Cross Blue Shield, HMO, Workers Comp | 6.5 | 5.7 |
|  | Medicare | 4.4 | 4.4 |
|  | Medicaid | 47.8 | 48.7 |
|  | Other Govt Payments | 24.5 | 22.1 |
|  | No Charge (Free, Charity, Special Research, Teaching) | 3.1 | 4.3 |
|  | Other | 6.4 | 6.7 |
| Premature Treatment Exit | No | 100 | 0 |
|  | Yes | 0 | 100 |

